# Equity gaps in the genetic diagnosis of monogenic cardiomyopathy: an intersectional analysis of race and gender in the All of Us Research Program

**DOI:** 10.64898/2026.11.23.26368743

**Authors:** Emilie T. Théberge, Spencer J. Anderson, Anna M. Lehman

## Abstract

**Purpose:** Diagnostic yield from genetic testing in cardiomyopathy depends on reference data and access to care, which differ by race and gender, yet are rarely studied jointly.

**Methods:** We analyzed 414,830 genome-sequenced *All of Us* participants for predicted deleterious variants (PDVs) in 58 cardiomyopathy genes, comparing ClinVar classifications by race and modeling cardiac outcomes against matched controls and, within heterozygotes, by race and gender. Among *GLA* or *TTR* pathogenic/likely pathogenic (P/LP) carriers with cardiomyopathy, we explored Fabry disease or amyloidosis diagnosis codes by race, gender, income and education.

**Results:** PDVs in White participants were more often P/LP (13.9% vs 12.0%, p<0.001), more so among heterozygotes with cardiomyopathy (29.6% vs 22.1%, p=0.004). Cardiomyopathy odds rose from missense to loss-of-function PDVs (OR 1.12 to 1.79). Twenty of 131 *GLA*/*TTR* P/LP carriers with cardiomyopathy had a syndromic code; White women had higher odds than non-White women (OR 6.50, 95% CI 1.35–31.34). Syndromic codes were nominally associated with higher income and education, and 47 of 48 carriers with income <$25k were non-White.

**Conclusion:** Variant classification and syndromic diagnosis are less complete for non-White carriers, particularly women. Equitable precision cardiology requires more diverse reference data and analyses that treat race and gender as effect modifiers.

## INTRODUCTION

Genetic testing in cardiovascular medicine has become increasingly accessible and cost-effective for clinicians to integrate into care pathways, directly informing medical management, cascade screening of relatives, and risk stratification for major outcomes.^1^ These benefits are notable in hereditary cardiomyopathies, a common group of diseases affecting the structure and/or function of the heart muscle that can lead to premature death from heart failure or cardiac arrest. Monogenic causes are identified in approximately 30 to 40% of (dilated) nonischemic cardiomyopathy (NICM)^2^ and 60% of hypertrophic cardiomyopathy (HCM) patients.^3^ Monogenic causes of cardiomyopathy in non-sarcomeric genes also expressed in extracardiac tissues (i.e., syndromic disease) illustrate how genetic testing can directly impact medical management: for example, characterizing pathogenic variants in the *GLA* or *TTR* genes that cause Fabry disease (FD, OMIM #301500) and hereditary transthyretin amyloidosis (ATTR, OMIM #105210), respectively, guides the decision to initiate disease-specific therapies.^4,5^

However, as the volume of genetic testing being conducted in clinical labs and for research increases, patients identifying as White or of majority European ancestry are disproportionately represented among those utilizing genetic testing, and by extension, the benefits of tailored care.^6^ Under-representation of non-European participants in allele frequency databases used to classify variants impacts diagnostic uncertainty: among 2011-2021 ClinVar reclassifications in cardiomyopathy genes, most clinically meaningful changes moved towards uncertain or conflicting significance, with about a third of pathogenic/likely pathogenic downgrades reflecting higher minor allele frequencies newly recognized in non-European populations.^7^

Expression of monogenic variants can be modulated by modifiable risk factors such as hypertension and diabetes,^8^ and their detection by social determinants of health (SDOHs). Studies have demonstrated unequal access to diagnosis and treatment observed for racialized populations, women, and people with lower income and education.^9,10^ Further, penetrance of cardiomyopathy variants also differ by sex and gender. Sex differences are often attributed to estrogen’s effects on genes influencing cardiomyocyte hypertrophy, contractility and electrophysiology,^11^ whereas gendered differences are exemplified by how symptoms are interpreted and triaged, which may delay or miss diagnoses in women, who present later and with greater symptom burden.^10^ In statistical modelling, adjusting for sex or gender obscures these differences: de Marvao *et al.* demonstrate divergent sex effects in HCM using imaging phenotypes in the UK Biobank and argue that sex be modeled as an effect modifier, rather than adjusted for, when analyzing penetrance for HCM.^12^

Few studies have investigated the intersection of both sex or gender and race in monogenic cardiomyopathy. Using All of Us data, we asked: 1) Do ClinVar classifications of predicted deleterious variants (PDVs) in cardiomyopathy genes differ by race? 2) Do odds of cardiomyopathy, heart failure and cardiac arrest differ by PDV functional consequence, and within carriers, by ClinVar classification, race and gender? 3) Among GLA or TTR P/LP carriers with cardiomyopathy, do odds of a syndromic (FD or ATTR) diagnosis code differ by race, gender and SDOHs?”

## MATERIALS AND METHODS

### Study Cohort and Variables of Interest

The All of Us (AoU) Research Program version 8 controlled-tier dataset were analyzed, which is a program that prioritized the recruitment of groups historically under-represented in biomedical research.^13^ Variant classification analyses included all participants with short-read genome sequencing (GS; n=414,830). Phenotype analyses were restricted to those with ≥2 International Classification of Diseases 10^th^ edition (ICD-10) codes, and complete self-reported gender identity, sex assigned at birth and race (n=329,707). Full methods are detailed in the **Supplemental Methods**.

Participants were identified if they carried at least one predicted deleterious variant (“PDV”) within 58 cardiomyopathy-associated genes annotated as autosomal dominant, semidominant or X-linked disease as listed in the 2023 European Society of Cardiology guidelines for the Management of Cardiomyopathies.^14^ Genes were further selected if they had ClinGen evidence of disease association and ≥1 pathogenic or likely pathogenic (P/LP) variant in AoU.

A PDV was defined as a missense variant with a REVEL^15^ score over 0.8 or a predicted loss-of-function (LoF) variant (frameshift, nonsense, start-or stop-loss, or canonical splice-site; Illumina Nirvana 3.18), with allele frequency <0.05 overall and in every AoU population. *TTN* PDVs were limited to LoF variants in the A-band of the cardiac N2BA/N2B transcripts. Classifications were taken from the most recently evaluated ClinVar submission for each variant as of October 11, 2025 (**Supplemental Methods**) and grouped as pathogenic or likely pathogenic (“P/LP”), variant of uncertain significance (“VUS”), or “Not in ClinVar” (no P/LP, VUS or benign/likely benign classification); benign or likely benign variants were excluded. Participants are described as carriers where the group is defined by a P/LP PDV, and otherwise as heterozygotes (including hemizygous men for X-linked genes). *GLA/TTR* analyses included all P/LP variants regardless of REVEL or LoF status.

Demographics and SDOH responses were self-reported, and predicted majority genetic ancestry was derived from GS data (definitions in **Supplemental Table 1**). Participants with congruent self-reported gender and sex assigned at birth were analyzed as cisgender women and men (“women” and “men”). Race was analyzed in its seven AoU labels and as a binary of “White” versus all other responses, including “More than one population” and “None of these” (“Non-White”). Age was derived using the year of birth relative to the year 2025. Education and annual household income are reported SDOH variables. Employment, health insurance, home ownership, marital status and country of birth were also examined among *GLA/TTR* P/LP carriers, but showed no significant associations and are not reported, per the AoU Data and Statistics Dissemination Policy.

Clinical phenotypes of interest were defined using AoU-harmonized SNOMED concepts mapping to ICD-10 codes from linked electronic health record (EHR) data, with inclusions and exclusions made via physician adjudication. Primary phenotypes were cardiomyopathy, defined broadly (all codes under the SNOMED “cardiomyopathy” concept) and narrowly as NICM (excluding ischemic etiologies and concurrent coronary artery disease), heart failure (HF) and cardiac arrest (CA). Eight cardiovascular risk factors, two concurrent cardiac phenotypes that increase risk of the primary phenotype, and syndromic codes for FD and amyloidosis (excluding non-ATTR amyloidosis) were also extracted (**Supplemental Table 2**).

### Statistical Analyses

Categorical variables were compared with χ² tests excluding missing responses; in descriptive summaries, missing education (<5%) was excluded and missing income retained as a category. PDV heterozygotes with EHR data were matched 1:4 to participants without a PDV on age, sex and predicted ancestry (R “MatchIt” package, v.4.7.2), and logistic regression estimated odds ratios (ORs, 95% CI) for each phenotype by PDV type. Within single-PDV heterozygotes, odds of cardiomyopathy were modeled by binary race, binary gender and a four-level race-by-gender variable (reference: White women), stratified by ClinVar classification and adjusted for age. Within *GLA/TTR* P/LP carriers with cardiomyopathy, odds of a syndromic code were modeled by binary race and binary gender (each adjusted for age) and by a four-level race-by-gender variable (unadjusted, given small cells). *GLA* and *TTR* carriers were pooled because syndromic codes were few in each gene, with a *TTR*-only sensitivity analysis (108/131 carriers), which also avoids combining X-linked (hemizygous) and autosomal inheritance in gender comparisons. Comparisons between syndromic and non-syndromic groups in the *GLA/TTR* analyses used a Bonferroni threshold of 0.05/20 = 0.0025.

## RESULTS

### PDV Heterozygotes and Race

Among 414,830 participants with GS data, 44,138 (10.6%) had at least 1 PDV (41,241 with one, 2,897 with at least 2), with 47,469 unique PDVs represented. The racial composition of PDV heterozygotes resembled that of the GS cohort (White 50.6% vs. 54.0%; Black or African American 19.5% vs. 17.5%; **Supplemental Table 3**). P/LP proportions varied widely among PDVs identified in the 58 genes analyzed, from 58.0% of unique PDVs in PKP2 to <3% in *RYR2*, *MYH6* and *MIB1* (**Supplemental Table 4**).

Overall, 6,169 of 47,469 PDVs (13.0%) were annotated as P/LP. P/LP proportions were lowest among Native Hawaiian or Other Pacific Islander (5.1%) and Middle Eastern or North African participants (7.3%), followed by Black or African American (11.7%), American Indian or Alaska Native (12.2%) and Asian participants (12.4%) (**Figure 1A**). PDVs in White participants were more often P/LP than in non-White participants (13.9% vs 12.0%, p<0.001; **Figure 1B**).

**Figure 1.**
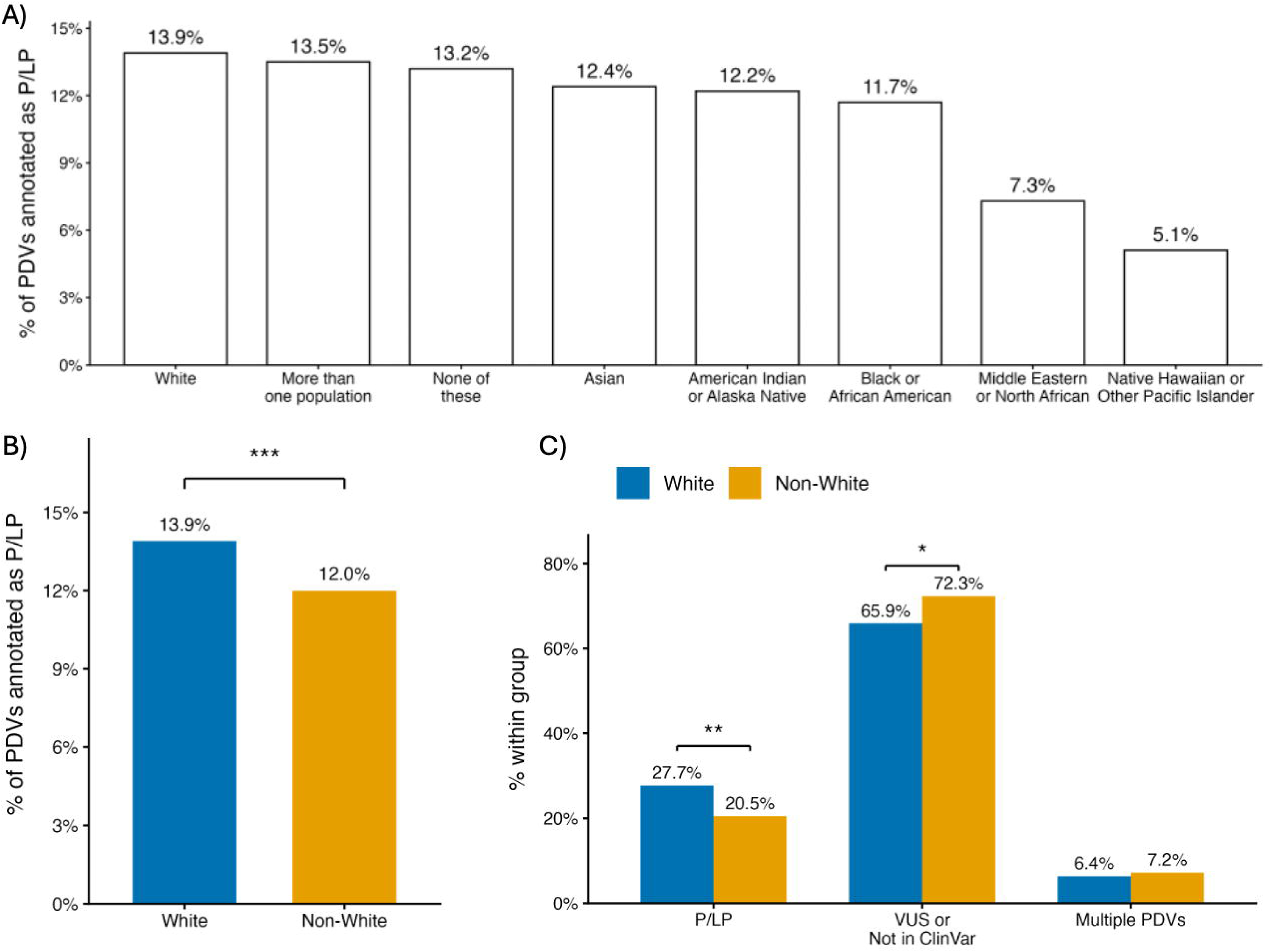
Proportions of predicted deleterious variants’ (PDV) annotations as pathogenic or likely pathogenic (P/LP), variant of uncertain significance (VUS) or not in ClinVar. A) Proportions of P/LP annotations among self-identified race categories among all PDVs (n=47,469). B) Proportions of P/LP annotated among PDVs by binary race (White versus non-White) of participants with a reported race (n=38,657 PDVs; Supplemental Table 3). C) Proportions between White and non-White PDV heterozygotes with cardiomyopathy (broad definition, n=1,462) by P/LP annotation or VUS/Not in ClinVar annotation (participants with single PDVs) or multiple PDVs. χ² test ***: p<0.001. **: p<0.01. *: p<0.05

Among 1,462 participants with cardiomyopathy (broad definition) and complete race data, 1,365 carried a single PDV and 97 multiple PDVs, and the proportion with multiple PDVs did not differ by race (7.2% vs 6.4%, p=0.64). Of all PDV carriers with cardiomyopathy, non-White participants less often carried a P/LP variant (20.5% vs 27.7%, p=0.003) and more often a VUS or Not in ClinVar variant (72.3% vs 65.9%, p=0.015; **Figure 1C, Supplemental Table 5**).

### PDVs and Cardiac Phenotypes

Relative to matched controls (n=110,992), odds of most cardiac phenotypes among PDV heterozygotes with EHR data (n=27,748) rose from single missense to multiple to single LoF PDVs, with the largest effects observed for nonischemic cardiomyopathy (**Figure 2; Supplemental Tables 6-7**). For broad cardiomyopathy, ORs were 1.12 (95% CI 1.04-1.21, p=0.002) for missense, 1.48 (1.21-1.79, p<0.001) for multiple and 1.79 (1.64-1.95, p<0.001) for LoF PDVs. The stricter NICM definition increased each OR relative to the broad definition (missense OR 1.20, 1.08-1.33, multiple PDVs OR 1.82, 1.43-2.33, LoF OR 2.05, 1.83-2.29; all p<0.001). Heart failure and the composite followed the same pattern at smaller magnitudes. Cardiac arrest was associated only with any PDV (OR 1.17, 1.02-1.35, p=0.027) and LoF PDVs (OR 1.38, 1.12-1.71, p=0.003).

**Figure 2.**
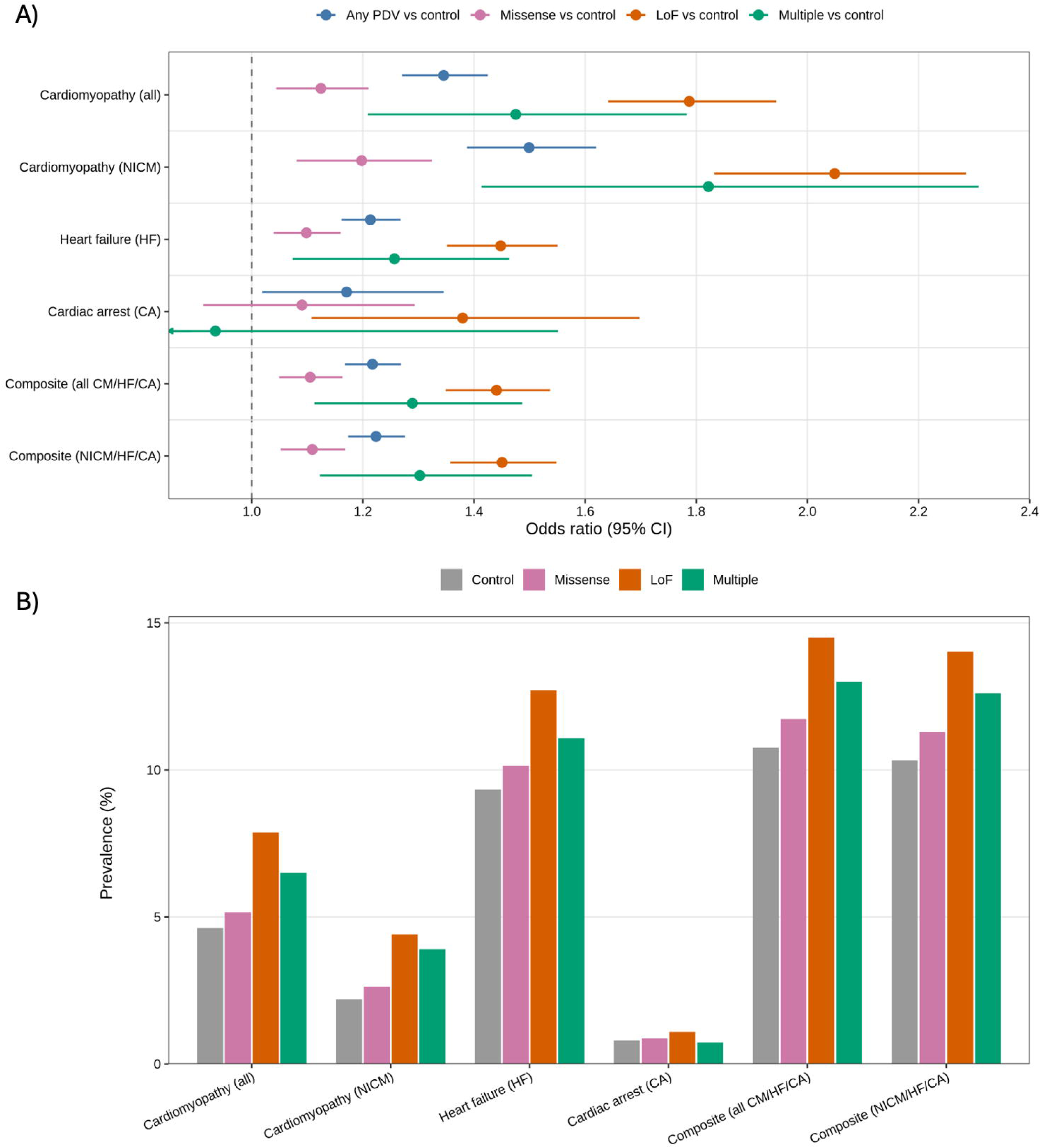
Association between predicted deleterious variant (PDV) heterozygote status and cardiac phenotypes compared to age-, sex-, and ancestry-matched controls with no PDV (1:4 carrier:control). A) Odds ratios (95% confidence intervals, CI) for each cardiac phenotype: cardiomyopathy (CM; broad or narrower nonischemic [NICM] definition), cardiac arrest (CA), heart failure (HF), and a composite of any of the three (CM/HF/CA). Four heterozygote comparisons are shown, each versus control: any PDV (all matched carriers, n=27,748), single missense PDV (REVEL>0.8) (n=17,310), single predicted loss-of-function (LoF) PDV (n=8,669), and multiple (participants with more than one PDV) (n=1,769). The missense and LoF groups are restricted to heterozygotes of a single PDV, so the missense, LoF, and multiple-PDV groups are mutually exclusive. Dashed line signifies OR=1. B) Prevalence (%) of each cardiac phenotype within matched controls and each mutually-exclusive PDV subgroup (single missense PDV, single LoF PDV, multiple PDVs).

Among single-PDV carriers, men had nearly twice the age-adjusted odds of cardiomyopathy (broad definition) compared to women for both P/LP (OR 1.83, 1.46-2.28, p<0.001) and VUS/Not in ClinVar heterozygotes (OR 1.96, 1.72-2.24, p<0.001; **Figure 3A, Supplemental Table 8**). Non-White VUS/Not in ClinVar heterozygotes had higher odds of cardiomyopathy than White heterozygotes (OR 1.31, 1.14-1.51, p<0.001), but significance was not observed by race for P/LP carriers. Relative to White women, White and non-White men with P/LP variants had higher odds of cardiomyopathy, but not non-White women (White men OR 1.74, 1.33-2.26, p<0.001; non-White men OR 1.82, 1.29-2.56, p<0.001) (**Figure 3B**). Non-White women had higher odds than White women only among VUS/Not in ClinVar heterozygotes (OR 1.40, 1.15-1.71, p<0.001).

**Figure 3.**
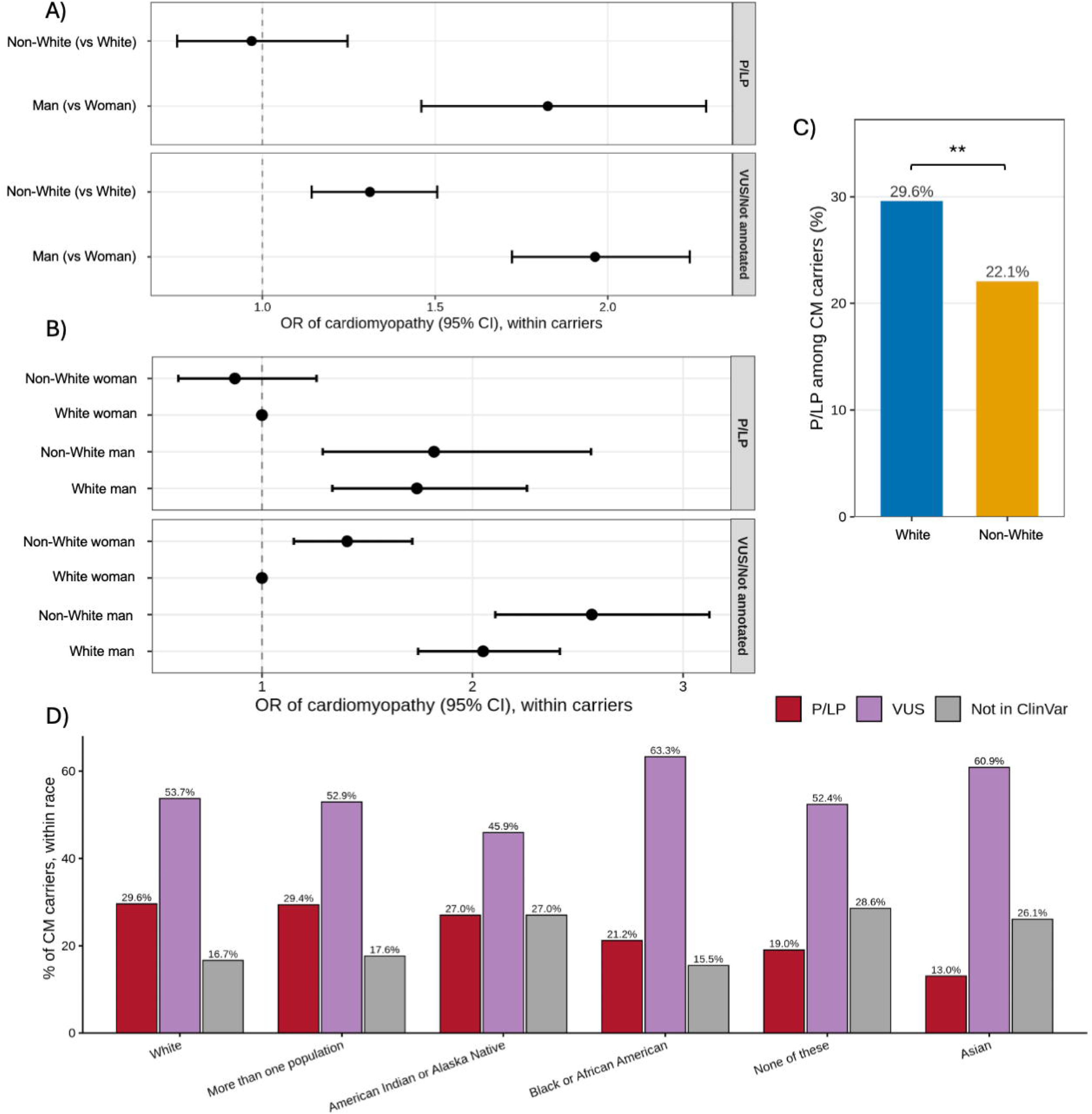
Cardiomyopathy (CM, broad definition) and variant classification by race and gender among single-PDV heterozygotes. A-B adjusted for age, dashed line signifies OR=1. A) Odds ratios (OR, with 95% confidence intervals, CI) for cardiomyopathy among PDV heterozygotes (n=41,241) by binary gender and binary race, stratified by whether the variant was classified as pathogenic/likely pathogenic (P/LP) or a variant of uncertain significance (VUS) or Not in ClinVar. B) OR (95% CI) for cardiomyopathy among PDV heterozygotes (n=41,241) in binary-race-by-binary-gender analyses. C) Proportion of variants annotated as P/LP among participants with cardiomyopathy (n=1,365), stratified by binary race. χ² test ** = p<0.01. D) Frequencies (%) of variants annotated as P/LP, a VUS, or Not in ClinVar among single-PDV heterozygotes with cardiomyopathy (n=1,365) stratified by race. For D), the total number of participants identifying as “Native Hawaiian or Other Pacific Islander” or “Middle Eastern or North African” were n<20, so were excluded from this figure.

### Syndromic Diagnostic Codes Among *GLA* & *TTR* P/LP Carriers

There were 2,220 carriers of a P/LP annotated variant in *GLA* (n=446) or *TTR* (n=1,774) who also had linked EHR data and complete gender and race data. Of these 2,220, only 36 had a syndromic diagnosis code of Fabry disease (n=7/446, 1.6%) or amyloidosis (n=29/1,774, 1.6%). Two *GLA* alleles accounted for most *GLA* carriers, p.(Ala143Thr) (n=202, 45.3%) and p.(Arg118Cys) (n=163, 36.5%), and one *TTR* allele for most *TTR* carriers, p.(Val142Ile) (legacy name p.Val122Ile; n=1,633, 92.1%) (**Supplemental Table 10**). Of the 2,220 carriers, 131 (5.9%) had broadly defined cardiomyopathy: 23 with *GLA* variants, 4 (17.4%) of whom had an FD code, and 108 with *TTR* variants, 16 (14.8%) of whom had an amyloidosis code. Eighteen of the 23 GLA carriers (78.3%) had p.(Ala143Thr) or p.(Arg118Cys), none with an FD code; 95 of the 108 TTR carriers (88.0%) had p.(Val142Ile).

Demographics, SDOHs, cardiovascular phenotypes and risk factors for the *GLA/TTR* P/LP carriers (n=2,220), carriers with cardiomyopathy (n=131), and the *TTR* P/LP carriers with cardiomyopathy (n=108) are summarized in **Supplemental Table 11**. Among *GLA/TTR* P/LP carriers with cardiomyopathy (n=131), the majority identified as Black or African American (76.3%), then White (19.1%), and a slight majority were women (51.9%). Carriers with cardiomyopathy (n=131) versus the all-carriers group (n=2,220) had more sleep apnea (51.1% vs 18.0%), chronic kidney disease (54.2% vs 14.2%), hypertension (86.3% vs 57.0%), valve disease (58.0% vs 12.0%), and myocardial infarction (38.2% vs 7.8%), as well as older age (77.1% vs 48.8% 60 or more years old). Among carriers with cardiomyopathy, only hypertension differed after Bonferroni correction: it was more common without a syndromic code (102/111, 91.9%) than with one (11/20, 55.0%; p=4.9 x 10^−5^), with a similar pattern among *TTR* carriers (92.4% vs 62.5%, p=0.003).

Neither race nor gender alone was associated with a syndromic code among pooled carriers (**Supplemental Table 12**). In race-by-gender analyses, White women with cardiomyopathy had higher odds of a syndromic code than non-White women (OR 6.50, 1.35-31.34; 4/12 vs 4/56) (**Figure 4A)**. Among *TTR* carriers only, White participants had higher odds of an amyloidosis code than non-White participants (OR 8.86, 1.78-49.13, p=0.008), as did White women (OR 16.33, 1.67-159.75, p=0.02) and White men (OR 32.67, 2.27-471.01, p=0.01) relative to non-White women (**Figure 4B**).

**Figure 4.**
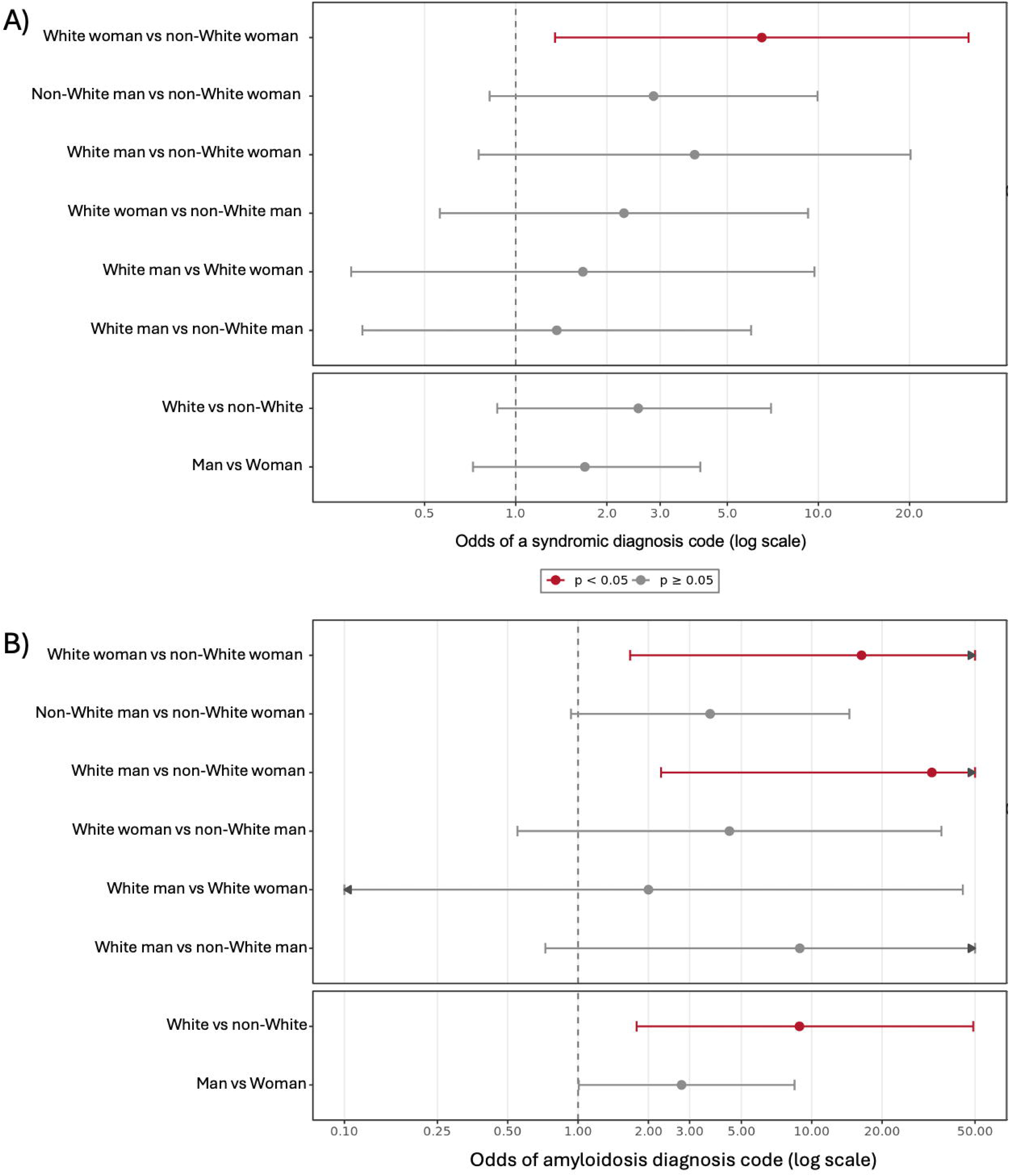
Forest plot of odds ratios (ORs, log scale, with 95% confidence intervals, CIs) of having a syndromic diagnosis code among participants with a cardiomyopathy code and A) a *GLA* or *TTR* pathogenic or likely pathogenic (P/LP) variant (n=131), or B) only a *TTR* P/LP variant (n=108; amyloidosis code). Upper rows: race-by-gender contrasts; lower rows: binary race or gender alone. Dashed line signifies OR=1. Arrowheads denote CIs that extend beyond the axis; red indicates p<0.05 (ORs unadjusted for multiple comparisons).

Before correction for multiple testing, education (p=0.005) and income (p=0.0065) differed by syndromic diagnosis status among *GLA/TTR* P/LP carriers with cardiomyopathy (n=131; **Figure 5A-B, Supplemental Table 11**). Participants with a syndromic diagnosis more often reported an annual household income ≥$100k (25.0% vs. 6.3%) and less often <$25k (15.0% vs. 40.5%), and more often reported some college or higher (95.0% vs. 59.4% of respondents). Income, but not education (p=0.12), differed by race and gender (p=0.00066; **Figure 5C-D, Supplemental Table 14**). Of the 48 participants reporting <$25k, 47 (98%) were non-White (25 women, 22 men). A similar, non-significant pattern was seen for education: 40 of the 44 participants (91%) reporting high school or less were non-White.

**Figure 5.**
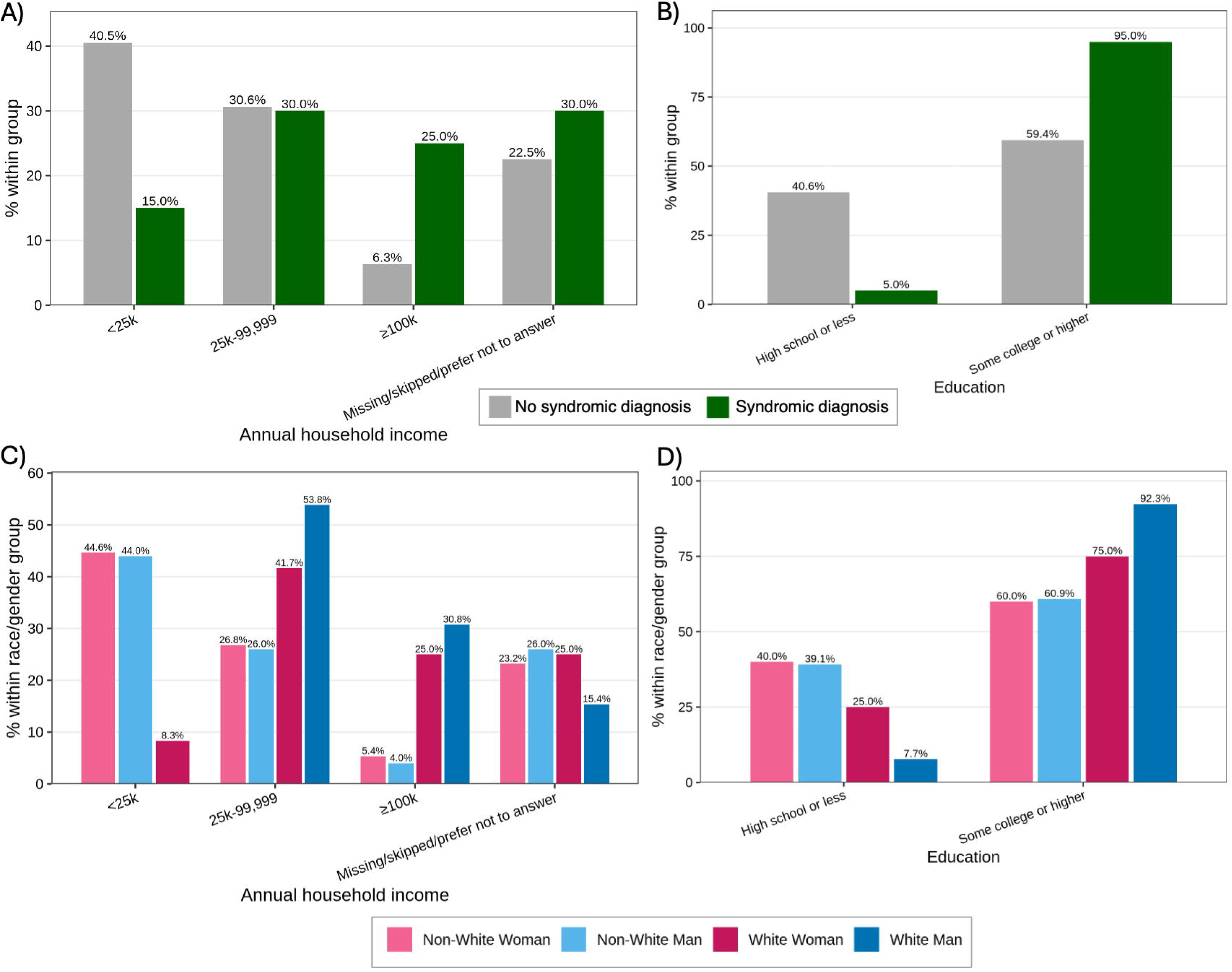
Annual household income (USD) and highest level of education attained among *GLA* or *TTR* P/LP variant carriers with cardiomyopathy (n=131), shown as percentages within each group. A) Income by syndromic diagnosis code status (with, n=20; without, n=111). B) Education by syndromic diagnosis code status (with, n=20; without, n=106). C) Income by race and gender: non-White women (n=56), non-White men (n=50), White women (n=12), White men (n=13). D) Education by race and gender: non-White women (n=55), non-White men (n=46), White women (n=12), White men (n=13). Income includes a missing/skipped/prefer-not-to-answer category; participants with missing education (n=5) were excluded from B and D.

## DISCUSSION

This work extends race- and gender-equity research in monogenic cardiomyopathy with an intersectional design, stratifying by both race and gender in the highly diverse AoU biobank. We observed that White PDV heterozygotes were more likely to have a P/LP annotation, across all PDV heterozygotes (13.9% vs 12.0%) and among those with cardiomyopathy (29.6% vs. 22.1%). Applying the latter percentages to an estimated 0.7-1.6 million Americans with heritable cardiomyopathy (using population prevalence estimates of 1:220 to 1:500,^2,3^ a 7.5-percentage-point difference in P/LP annotations corresponds to the order of 52,500-120,000 individuals with potentially reduced diagnostic clarity from genetic testing results returning uncertain or unannotated results. Closing this annotation accuracy gap is instrumental for mainstreaming precision medicine into primary and disease-specific care pathways for all.^16^

Our results agree with previous evidence of racial disparities in cardiomyopathy variant annotations in AoU,^17^ which did not stratify analyses by sex or gender. Race here is a self-identified social category and not a proxy for genetic ancestry, the basis on which reference databases such as gnomAD are built.^18^ We demonstrated significant differences in the frequency of PDV annotations as P/LP by race, highlighting the need to increase recruitment of under-represented populations in genetics research and to increase clinical genetic testing access to gain the evidence needed to classify more VUS as pathogenic or benign. Meaningful engagement and respectful partnership are essential when approaching marginalized communities about health research. Historical misuse of genetic data and misrepresentation of how it would be used, including its commodification,^19^ have fostered a warranted mistrust of the Western and colonial institutions that hold these data. There are particular considerations for American Indian or Alaska Native, Black or African American communities, and Native Hawaiian or Other Pacific Islander populations (discussed by Fox^19^, Arbour & Cook^21^, Scherr *et al.*^21^ and Taitingfong *et al.*^22^).

Historically, intersectional race-by-gender analyses have been underpowered in genomic cohorts lacking both ancestral diversity and gender parity, resulting in authors adjusting for one variable rather than stratifying across both, which can obscure genuine effect modification.^23^ By leveraging the AoU biobank, which has sufficient scale to cross these axes directly for rare monogenic disease, we found that the elevated odds of cardiac phenotypes among PDV heterozygotes (most pronounced for LoF variants and cardiomyopathy) were driven predominantly by gender rather than race: men had nearly twice the odds of cardiomyopathy as women across both P/LP and VUS/unannotated strata. Moreover, among heterozygotes with a VUS/Not in ClinVar variant, non-White women had significantly higher odds of cardiomyopathy compared to White women, suggesting that there may be an enrichment for as-of-yet pathogenic variants within the unannotated group. These results are consistent with well-documented sex differences in the penetrance and expressivity of genetic cardiomyopathies, in which men more often and more severely express disease.^10^ The magnitude and consistency of this effect argues that sex should be modeled as an effect modifier, not just a covariate, in penetrance and risk estimates for carriers of pathogenic variants in cardiomyopathy genes.

While we propose that the unannotated group may conceal unrecognized pathogenicity, the incomplete penetrance of two recurrent alleles in much of the carrier and cardiomyopathy cohorts (*GLA* p.(Ala143Thr) and *TTR* p.(Val142Ile)) warrants specific attention based on the low prevalence of cardiomyopathy in these P/LP carriers relative to controls without a PDV or P/LP variant. The pathogenicity of *GLA* p.(Ala143Thr) is subject to ongoing debate, with some proposing that it be classified as a “variant of varying clinical consequence”^24^ or that it be associated with “mild GLA insufficiency” rather than true Fabry disease.^25^ Although p.(Ala143Thr) accounted for 9 of the 23 *GLA* carriers with cardiomyopathy (39.1%), only 4.5% of its 202 carriers had cardiomyopathy, similar to the 4.6% prevalence among matched non-carriers (**Supplemental Table 7**), and none had an FD code. This is consistent with low penetrance of a variant of debated pathogenicity.^26^ The most common *TTR* variant, p.(Val142Ile), accounted for 88% of cardiomyopathy among *TTR* P/LP carriers but affected only 5.8% of its 1,633 carriers. There was a significantly high rate (91.9%) of hypertension among cardiomyopathy *GLA/TTR* P/LP carriers without a syndromic diagnosis code; these participants may have been presumed to have had hypertensive cardiomyopathy, and not investigated further for monogenic disease like FD or ATTR, which may explain the absence of syndromic codes. These findings add to evidence of underdiagnosis of these syndromic phenocopies in other cardiomyopathy cohorts.^27,28^

Lower income and education were nominally associated with lacking a syndromic diagnosis code, with non-White women and non-White men comprising nearly all of the lowest-income stratum. As social determinants can interact with disease risk as indicators of access or capacity to engage with healthcare institutions,^9^ these results are consistent with an equity gap in access to genetic testing. Increasing health literacy and reducing the economic barrier to genetic testing and access to disease-modifying therapy are critical for reducing the disease burden in these groups. Besides the moral argument for providing equitable healthcare access to all populations, reducing inequities leads to economic benefits: health inequities, through premature death, excess medical costs and lost productivity, cost the USA an estimated USD $451 billion in 2018.^29^

Variable, often low penetrance of common alleles complicates the timing to initiate costly FD- or ATTR-specific therapy. Still, carriers with established cardiomyopathy are the likeliest treatment candidates, and the absence of syndromic codes in most of the 131 suggests that at least some represent missed genetic diagnoses. Among *TTR* P/LP carriers with cardiomyopathy, women and men were nearly equally represented (51.9% vs 48.1%), yet men held 11 of the 16 amyloidosis codes. This contrasts with other ATTR clinical cohorts, which are 80-90% male,^30^ suggesting under-recognition in women. Our intersectional results point to non-White women as warranting particular attention in diagnosis, registry recruitment and efforts against systemic bias, since under-enrollment of women in cardiovascular research is compounded by racial inequity, particularly for Black or African American women. Paquin *et al.* recommend institutions dedicated to underserved and under-represented populations and lowering financial barriers to access may assist in these efforts.^31^

There are several limitations of this study inherent to the dataset used, and *in silico* modeling relied on to predict deleteriousness of variants within the genes of interest. The use of ICD-10 codes for phenotype ascertainment, in the absence of more granular echocardiogram or cardiac MRI data, can lead to both false negatives (undiagnosed) and false positives (miscoded or misdiagnosed) within the dataset. The linked EHR data did not record who was genetically tested. The decision to study FD and ATTR as the “genetic diagnoses” of interest relied on the assumption that these were unlikely to be miscoded given their specificity. In contrast, studying individuals with ICD-10 codes for HCM (which FD and ATTR are phenocopies of) risks conflating true genetic HCM with secondary (non-hereditary) left ventricular hypertrophy driven by hypertension, aortic stenosis, and/or metabolic mimics, leading to false positives and misclassification, as exemplified by Nielsen *et al*.^32^ Moreover, there is also likely an under-coding of true familial HCM patients with potentially more general “cardiomyopathy” codes, as posed by researchers interrogating HCM in the UK biobank.^33^ This further justified the use of a broad, general “cardiomyopathy” code in our analyses. For the variant predictions, we recognize the inherent limitations with using an arbitrary REVEL cutoff of 0.8 as a missense threshold,^34^ which was chosen to reduce false positives; there are pathogenic variants that have REVEL scores under 0.8, exemplified by the variants present in our P/LP *GLA* and *TTR* results (**Supplemental Table 10**), and benign variants with scores over 0.8.

In conclusion, we show that heterozygotes of PDVs from non-White groups are less likely to receive a pathogenic annotation, and among *GLA/TTR* P/LP carriers with cardiomyopathy, non-White women were least likely to have a concurrent FD or ATTR diagnosis code. These gaps more plausibly reflect inequities in variant curation and access to genetic diagnosis by race rather than differences in underlying biology. Furthermore, the observation that the lowest-income *GLA/TTR* P/LP carriers were almost entirely non-White and the least likely to hold a syndromic diagnosis code points to gaps in healthcare access, not genomics alone. Closing these gaps will require both the deliberate recruitment of under-represented populations into genomic databases and intersectional analytic approaches that treat sex or gender and race as effect modifiers rather than covariates, so that the benefits of precision cardiovascular medicine can serve everyone.

## Declaration of AI and AI-assisted technologies in the writing process

During the preparation of this work, the first author used Claude (Anthropic) to assist in writing code for analyses, as well as proofreading manuscript contents for clarity and accuracy. After using this tool/service, all authors reviewed and edited the content as needed and take full responsibility for the content of the publication.

## Data Availability

This study used data from the All of Us Research Program’s Controlled Tier Dataset v8 (C2024Q3R3), available to authorized users on the Researcher Workbench (https://www.researchallofus.org). Access requires institutional registration, a Data Use and Registration Agreement, and completion of All of Us research ethics training; participant-level data cannot be shared by the authors under these terms. We requested and received an exception to the All of Us Data and Statistics Dissemination Policy from the All of Us Resource Access Board to report counts under 20. Code used for analyses is available from the corresponding author upon reasonable request.

## Acknowledgements

We gratefully acknowledge All of Us participants for their contributions, without whom this research would not have been possible. We also thank the National Institutes of Health’s All of Us Research Program for making available the participant data examined in this study.

## Author Contributions

Conceptualization: E.T.T., A.M.L.; Data curation: E.T.T., S.J.A.; Formal analysis: E.T.T., S.J.A.; Funding acquisition: A.M.L.; Investigation: E.T.T.; Methodology: E.T.T.; Software: E.T.T., S.J.A.; Supervision: A.M.L.; Visualization: E.T.T.; Writing-original draft: E.T.T.; Writing-review & editing: E.T.T., S.J.A., A.M.L.

## Ethics Declaration

This study was approved by the University of British Columbia Clinical Research Ethics Board (H23-00216). It used de-identified data from the All of Us Research Program (https://www.researchallofus.org), whose participants provided informed consent under protocols approved by the All of Us Institutional Review Board; no additional consent was required for this secondary analysis. The study adhered to the principles of the Declaration of Helsinki.

