## Supplemental Methods for "Equity gaps in the genetic diagnosis of monogenic cardiomyopathy: an intersectional analysis of race and gender in the All of Us Research Program"

### 1. Defining Variables in All of Us

#### 1.1. Demographics

Demographic variables were extracted from the survey “The Basics”: Self-reported gender identity, sex assigned at birth, age, and self-reported race. Predicted majority genetic ancestry derived from genome sequencing data, were reported alongside demographics. Demographic variables and labels are summarized in **Supplemental** **Table 1**.

##### 1.1.1 Sex and Gender

Sex and gender are highly correlated, but confer different risk for cardiovascular disease through distinct biological mechanisms (e.g. sex-specific gene expression, gonadal hormone levels) and gender identity (e.g. differential risk via psychosocial factors).^1^ As the AoU authors describe purposeful sampling of under-represented groups in biomedical research^2^, including sex and gender identity minorities, gender identity and sex assigned at birth could not be assumed to be congruent. Therefore, a new variable was made to capture responses from both gender and sex variables, with options: cisgender female, cisgender male, and an “other” category for non-congruent answers. For example, a participant who responded “female” sex at birth but skipped answering gender identity cannot be assumed to be a cisgender woman. This choice to group the variables as such was made after reviewing Canadian Institutes of Health Research guidance for sex and gender methodology, while in the context of this study analyzing cardiovascular disease risk, for which biological sex of male (46XY) and female (46XX) and cisgender risk are highly correlated.^3^ There is emerging research on how cardiovascular risk differs by gender identity^4^ and by biological sex.^5^ However, given the low proportion of gender-sex incongruence in the AoU dataset, these analyses opted to use a binary of cisgender males versus cisgender females (hereafter referred to as “men” and “women”, respectively) to ensure adequate statistical power for downstream analyses within the subset of monogenic cardiomyopathy variant carriers.

##### 1.1.2 Age

Age for all participants was provided in the form of a year at birth, with all months and days of birth set to June 15. An age at analysis start was derived from subtracting this year of birth from 2025, and a new grouped variable also classifying them into three categories: under 40, 40-59, and 60 or over. For participants with any major cardiac phenotype or genetic diagnosis of interest, an “age at earliest diagnosis” value for each phenotype was derived from the earliest date of the phenotype code occurring in the EHR and also grouped into the same 3 age categories for sub-analyses.

The lower threshold of 40 years has been used for genetic testing in multiple guidelines, including the United Health Care Genetic Testing for Cardiac Disease guidelines in the context of patients with sudden cardiac arrest without known structural heart disease.^6^ Recent Canadian guidelines for cardiogenetic testing describe a high rate of monogenic findings in sudden cardiac death under 35 years, but this is not stated explicitly as a guideline.^7^ The choice of 40-59 years to represent “middle age” is the same range as defined by the Encyclopedia Britannica.^8^ Furthermore, these two decades represent a significant transition to increase in cardiovascular risk for many women due to menopause, with this specific age range (40-59) supported by a Scientific Statement from the American Heart Association^9^ regarding the menopause transition and cardiovascular disease risk. Lastly, a recent report in the Lancet used these same age categories (<40, 40-59, 60+) when reporting trends in CVD outcomes in the US by income and education.^10^

A limitation with deriving age at diagnoses with these data is that it isn’t possible to verify if this was the true age at first diagnosis, as it’s possible that a diagnosis preceded the digitalization of the health records. The key milestone for EHR adoption in the US was in 2009, when the Health Information Technology for Economic and Clinical Health Act allocated $19.2 billion in funding to encourage healthcare providers to adopt EHRs through Medicare and Medicaid.^11^ The rapid uptake resulted in 84% of non-federal acute care hospitals having adopted at least a basic EHR system, and 96% possessed a certified EHR technology.^12,13^ In 2015, hospital basic EHR adoption rates were above 65% across all states.^12,13^

##### 1.1.3 Race

Historically, the social constructs of race and ethnicity have been used to generalize groups of people based on physical characteristics (skin, hair, eye colour) and cultural factors such as language, religion and shared nationalities.^14^ The options for self-reported race presented to participants were the following: “American Indian or Alaska Native”, “Asian”, “Black or African American”, “Middle Eastern or North African”, “White”, “More than one population”, and “None of these”. It is important to underscore that these are socially constructed, subjective terms, that were historically used as damaging constructs to establish racial hierarchies and justify discrimination. Following joint guidance issued across major genetics and genomics journals^15^, population descriptors such as race and geographic origin should no longer be used as proxies for genetic ancestry groups in genomic science. Thus, correlations between race and predicted genetic ancestry were not done.

Proportions of PDVs annotated as P/LP were compared between binary race (“White” versus “Non-White”) and binary majority genetic ancestry (“eur” versus “noneur”). Definitions of labels in each of these groups are expanded on in **Supplemental Table 1**. Relative differences were calculated using the formula: [ (White P/LP %) - (Non-White P/LP %) ] / (Non-White P/LP %) for race, and [ (eur P/LP %) - (non-eur P/LP %) ] / (non-eur P/LP %). Chi-squared tests of significance were used to determine if the proportional differences by binary category were significantly different.

#### 1.2 GS data and Genetic Ancestry

Short-read genome sequencing (GS) was conducted on 414,830 participants in the All of Us controlled tier dataset (version 8, released February 3, 2025). Full details of the quality control (QC) pipeline are detailed extensively in the report “All of Us Curated Data Repository (CDR) release C2024Q3R3”.^16^ In brief, samples were constructed using PCR Free Kapa HyperPrep, sequenced on NovaSeq 6000 machines, and processed using DRAGEN v.3.7.8. Single sample QC was followed per Illumina protocols described previously^17^, including sex concordance, cross-individual contamination (pass: <3%), and coverage (≥30x mean coverage and ≥90% of bases at 20x coverage). GS SNP and INDEL joint callset QC was conducted on a joint callset, not individual samples, with a QC process similar to that of gnomAD v.3.1.^18^ This step helped identify noisy samples, artifacts, poor variant detection, flagging population outliers, calculation of allele frequencies, and flagged sample relatedness.

Genetic ancestry was inferred by measuring the genetic similarity of each participant to harmonized continental metadata labels from the Human Genome Diversity Project (HGDP)^19^ and the 1000 Genomes Project training data (N=3,942)^20^ for all GS samples. The population category names used for analyses were African (afr), Americas (amr), East Asian (eas), European (eur), Middle Eastern (mid) and South Asian (sas). AoU analysts trained a random forest classifier on the first 16 principal components of the HGDP and 1000 Genomes samples, onto which AoU participants were projected.

#### 1.3 Social Determinants of Health

Select social determinants of health (SDOHs) were chosen primarily based on the variables included in an All of Us-specific social deprivation index proposed by Gupta *et al.*^21^, as well as variables well-described in epidemiological literature to associate with cardiovascular health: Education level, employment status, health insurance status, home ownership, annual household income, marital status, and country of birth, summarized in **Supplemental Table 1**. All of these questions were asked in “The Basics” survey. Variables were condensed into binary and/or categories with fewer options to facilitate having sufficient power for statistical analyses, following literature-guided justification.

#### 1.4 EHR data

##### 1.4.1 Phenotype Extraction from EHR

AoU uses the Observational Medical Outcomes Partnership (OMOP) Common Data Model to store and standardize electronic health record (EHR), survey, and physical measurement data.^22^ EHR condition source vocabularies (i.e., International Classification of Diseases (ICD)-10 codes) are mapped to Systematized Nomenclature of Medicine (SNOMED) as the OMOP standard vocabulary for conditions. Each concept is identified by a standard “concept ID” (a unique OMOP integer) distinct from its source “concept code” (the native ICD-10 or SNOMED identifier); as multiple source codes map to a single concept ID, phenotypes were defined using standard SNOMED concept IDs to capture diagnoses irrespective of their original source vocabulary.

The version 8 dataset identifies 393,601 participants as having linked EHR data out of the total 633,534; however, there is a proportion of these 393,601 participants who do not have any EHR entries, defined as mapped ICD-10 codes per above. A brief literature review was conducted to identify justifiable thresholds for how many ICD-10 codes a participant should have to be included in this EHR-denominator participant pool. This step was needed to reduce the number of participants included in the background as potential “false negative EHR controls” of participants without any ICD-10 codes, despite being in the EHR-linked group.

Wei *et al.*^23^ evaluated ICD-code phenotyping against manual chart review across 10 diseases and found that requiring two or more ICD codes improved positive predictive value (0.84) compared with using ICD codes more loosely (0.06-0.71). Sulieman *et al.*^24^ reported that, of 203,813 participants with linked EHR data in AoU version 4 (314,994 total), 190,590 had ≥2 ICD-10 codes. The authors cite multiple phenotype-wide association studies^25, 26, 27^ as justification for this foundational convention of the use of repeated ICD codes improving phenotyping reliability. More recently, Felker *et al.*^28^ used a minimum of at least 1 ICD-10 code among AoU version 7 data. This filter resulted in 182,459 participants analyzed, out of the total 287,012 participants originally indicated as having EHR data linked (a 36.4% reduction) in v7.

##### 1.4.2 Cardiovascular Phenotypes and Risk Factors

Among the denominator of participants with at least 2 of any ICD-10 codes, specific cardiovascular phenotypes and relevant risk factors/risk modifiers were further subset and quantified. Cardiomyopathy, cardiac arrest and heart failure were chosen as primary cardiac phenotypes of interest for interrogating hypotheses of PDV carriers versus non-carriers due to their frequent association with the cardiomyopathy genes.

For cardiomyopathy, two approaches were taken when choosing the specificity of the included terms. First was a “catch-all” subset of all participants with any of the ICD-10 terms nested under the harmonized SNOMED code “cardiomyopathy”. Then, a more conservative subset was defined by removing cardiomyopathy terms with ischemia named within the cardiomyopathy, or implied ischemia based on if there was concurrent coronary artery disease.

The justification of including ischemic etiologies within the first pass is based on increasing evidence that monogenic variants in known cardiomyopathy disease genes have been associated with increased odds of cardiomyopathy pathology and worse outcomes developed post-ischemic insult.^29, 30, 31^ The sensitivity analysis excluding ischemic etiologies will distinguish the degree to which the inclusion of ischemic cardiomyopathy has on the odds ratios of PDVs associating with this outcome. ICD-10 codes included and excluded for each cardiac phenotype are summarized in **Supplemental Table 2**.

Risk factors that have been shown to be associated with increased risk of cardiomyopathy and/or heart failure incidence in epidemiological studies were also extracted, as summarized in **Supplemental Table 2**. The frequency of these risk factors/modifiers were reported within PDV carriers versus non-carriers among participants with any of the three main cardiac phenotypes of interest, then within participants with cardiomyopathy. Given the paucity of literature on intersectional reporting of cardiac outcomes and risk factors by sex or gender and race, hypothesis-generating counts of the cardiac outcomes and risk factors were further reported by binary gender and binary race (White vs non-White) within the PDV carriers vs non-PDV carriers with any of the 3 main cardiac phenotypes of interest, and in 4-group intersectional analysis (gender*race).

Frequency of the following risk factors (potential confounders) were reported to compare between PDV carriers and non-PDV carriers: alcohol use disorder, chronic kidney disease, diabetes mellitus, dyslipidemia, hypertension, obesity, obstructive sleep apnea, and smoking history. Smoking history was the only variable derived from a self report, coded from the question with yes/no/skip options: “Have you smoked at least 100 cigarettes in your entire life? (There are 20 cigarettes in a pack.)”, OMOP concept ID 1585857. The tabulated options were summarized as a binary “Ever” (response: Yes) versus “Never” (response: No). Hypertension can independently cause left ventricular hypertrophy, heart failure and arrhythmias, such that it is both a confounder as well as a potential effect modifier.

Chi-squared tests of significance were used to evaluate if there were differences in the frequency of these other variables between PDV carriers and non-carriers.

#### 1.5 Control group selection methods

The R package “MatchIt” was used to identify controls matched for age, sex, and majority predicted genetic ancestry to PDV carriers at a 1:4 carrier:control ratio. The marginal gain in statistical power is negligible beyond a 1:4 ratio. Efficiency improves substantially up to about 4 controls per case and then plateaus, with each additional control beyond four contributing under ~2% to the precision of the effect estimate. A 1:4 ratio therefore captures nearly all the available power while preserving a large, well-matched control pool.

### 2. Defining cardiomyopathy gene list and PDV criteria

#### 2.1 Gene List

The 2023 European Society of Cardiology (ESC) guidelines for the management of cardiomyopathies^32^ provide a table of 96 genes associated with monogenic syndromic and non-syndromic cardiomyopathies. Of these 96 genes, 83 have autosomal dominant (AD), semidominant (SD) or X-linked (XL) inheritance patterns, which were selected for the initial variant filtering analysis.

In the AoU research workbench, when subsetting the individual-level data contained in the Variant Dataset in the HAIL genomics format, the gene limits for variant selection were defined per GRCh38 build Ensembl 114 with gene boundaries ±5kb flanking the longest transcript. This range was used to match the stated limits of the genes annotated in the corresponding Variant Annotation Table documented by AoU.^33^ Only MANE Select or MANE Clinical Plus transcripts were included.

Following variant filtering, a *post hoc* analysis was conducted to narrow down the included genes (n=83) for analysis using more stringent criteria. First, genes were identified that had ClinGen evidence suggestive of disease, whereby they had at least one gene-disease relationship classified as “Definitive”, “Strong”, “Moderate”, or “Limited”. Genes that did not have entries in ClinGen, genes that were stated as having “No known disease association”, or genes that had annotations of “Disputed” or “Refuted” were excluded at this next step. Genes with multiple disease associations, such as “Definitive” evidence for DCM but “Disputed” evidence for HCM, were retained if at least one cardiomyopathy (or syndromic disease that can result in a cardiomyopathy) had suggestive evidence. Secondly, after predicted deleterious variants were identified, genes that did not have at least 1 variant annotated as P/LP were removed from downstream analyses.

The purpose of the post hoc analyses was not to capture ***all*** possible carriers of pathogenic variants in monogenic cardiomyopathy genes in the AoU cohort. Rather, the purpose was to evaluate equity gaps in variant annotations among people who carry predicted deleterious or pathogenic variants in the most common, robustly annotated genes annotated to cardiomyopathy that have a higher likelihood *a priori* of their variants having a pathogenic annotation. Genes annotated to AD, SD or XL inheritance with evidence of gene-disease association in ClinGen were chosen as the AoU data are not phased, so it would not be possible to distinguish if carriers of 2 variants in a gene annotated to autosomal recessive disease were compound heterozygotes or homozygous (i.e., in *trans*) or on the same chromosome (i.e., in *cis*).

#### 2.2 Defining Predicted Deleterious Variants

The Variant Annotation Table produced by AoU includes annotations produced by Illumina NIRVANA 3.18 software, including REVEL scores^34^, predicted loss-of-function (LoF) annotations, and ClinVar classifications of pathogenicity of benign (B), likely benign (LB), variant of uncertain significance (VUS), likely pathogenic (LP), pathogenic (P), or others, including “drug response” and “conflicting evidence”, including date stamps of the entry in ClinVar.

In the AoU Researcher Workbench, an original pipeline was developed for variant filtering specific to this thesis project. Variants were first filtered if they contained any of the following criteria: 1) Were annotated as pathogenic and/or likely pathogenic (“P/LP”); 2) had a REVEL score over 0.8; and/or 3) were predicted LoF. This REVEL cut-off score of 0.8 was decided as a relatively conservative threshold compared to ranges observed in other analyses to reduce the potential impact of statistical noise of potential non-pathogenic predicted deleterious variants (PDVs) with more moderate REVEL scores. This is supported by analyses conducted by Cubuk *et al.*^35^ who identified a higher positive likelihood ratio for their phenotypes of interest when using variants with REVEL scores ranging from 0.8-1 compared to <0.7. LoF was defined as *in silico* predicted frameshift, stop lost, stop gain, start lost, canonical splice donor site, or canonical splice acceptor site. A variant was classified as a PDV if it met REVEL or LoF criteria. Variants were filtered out if they had an allele frequency (AF) over 0.05 for any of the overall or subpopulation-specific AF as annotated per gnomAD (v.3.1) or within the AoU cohort. Per 2015 ACMG guidelines, a population AF over 0.05 is widely accepted as sufficient independent criteria to be considered a benign variant.^36^

Special considerations were applied to the *TTN* gene, the largest protein encoded by the human genome, while considering the variability in exact filtering criteria used by different authors who analyze *TTN* for rare variants that could cause cardiomyopathies following Mendelian inheritance patterns. Variants were retained if they were in the A-band of the adult cardiac *TTN* transcripts N2BA or N2B, as characterized by Roberts *et al.*^37^ and coordinates retrieved from the *TTN* entry in CardioDB.^38^ The A-band is the most constitutively expressed region, with minimal exon usage variation reported across major transcripts. In contrast, other regions contain exons with variable percentage spliced-in values. Further, variants were retained if they were LoF, as missense variants have less confident evidence of pathogenicity due to their high frequency in the general population, with estimates of up to 20% of the general population carrying a rare (AF<1%) *TTN* missense variant.^39,40^

ClinVar submission-level classifications (submission_summary file, downloaded October 11, 2025) were harmonized with the PDV dataset. For each variant, the classification from the submission with the most recent date of last evaluation (DateLastEvaluated) was used, and undated submissions were used only when no dated submission existed. This differs from ClinVar’s aggregate classification only where submitters disagree. Submission classifications other than P/LP, VUS or B/LB (e.g., “not provided”) were treated as Not in ClinVar. Variants with a ClinVar classification in the AoU annotation (2023) but no corresponding classification in the 2025 submission data were excluded. Variants whose most recent classification was B/LB were excluded, leaving three categories: P/LP, VUS, and Not in ClinVar. Recency was used on the premise that later evaluations draw on more cumulative evidence and current ACMG/AMP criteria: older ClinVar classifications are more often discordant with consensus,^41^ and interpretations predating current criteria are a leading source of inter-laboratory discordance.^42^ Because recency does not reflect the strength of evidence, a recent submission without assertion criteria, or from a single laboratory, could supersede an older, better-supported one, including an expert panel classification; this rule was chosen for reproducibility.
